# Microbial mechanisms underlying prebiotic-linked improvements in physical function and metabolism in knee osteoarthritis and obesity

**DOI:** 10.64898/2026.01.21.26344540

**Authors:** Weilan Wang, Rafael Fortuna, Shyamchand Mayengbam, Ruth A. Seerattan, Chunlong Mu, Jaqueline L. Rios, Nada Abughazaleh, Elnaz Vaghef Mehrabani, Erin W. Noye Tuplin, David A. Hart, Keith A. Sharkey, Walter Herzog, Raylene A. Reimer

## Abstract

**Background:** Knee osteoarthritis (OA) is a prevalent painful degenerative disease without effective disease-modifying drugs. The rising prevalence of comorbid obesity and knee OA underscores the urgent need for effective management to delay or prevent disease progression. In a recently completed randomized, placebo-controlled trial in adults with comorbid obesity (BMI >30 kg/m²) and unilateral or bilateral knee OA (Kellgren–Lawrence grade II–III), we were the first to demonstrate that a 6-month prebiotic intervention (16 g/day oligofructose-enriched inulin) significantly improved physical function and metabolic health.

**Methods:** To elucidate the underlying mechanisms, we incorporated metagenomics, metabolomics, and machine-learning-based multi-omics integration in 30 participants who completed baseline and at least one follow-up assessment and sample collection at months 3 and 6.

**Results:** Prebiotic supplementation reshaped gut microbial composition and function, increasing diet-derived carbohydrate availability, mitigating excessive host-glycan degradation and mucosal barrier disruption, reducing systemic inflammation and metabolic dysregulation, and ultimately improved physical performance and metabolic health. In a diet-induced obese rat model, prebiotic treatment reduced tibial cartilage degeneration and synovial membrane thickening, providing protection against OA onset and progression through a shared inflammatory pathway.

**Conclusions:** Our findings provide mechanistic evidence supporting the therapeutic potential of prebiotic supplementation as a conservative management in humans and as a preventive approach for obesity-related knee OA in a preclinical rat model, mediated through the gut-joint axis.

**Trial registration:** Clinicaltrials.govNCT04172688

## Introduction

Knee osteoarthritis (OA) is a painful degenerative joint disease characterized by the breakdown of articular cartilage, accompanied by osteophyte formation, pathological chondrocyte differentiation, and synovitis(1), affecting over 365 million people worldwide(2, 3). Despite its high prevalence, no effective disease-modifying OA drugs are currently available, leaving clinical management largely reliant on lifestyle modification, symptomatic analgesics and anti-inflammatory agents(4).

More than 50% of knee OA patients present with comorbid overweight or obesity(3, 5). Although the precise etiology is not yet clear, current consensus has shifted the pathophysiological role of obesity in knee OA from being solely attributed to excessive mechanical loading to systemic factors, particularly chronic systemic inflammation and impaired glucose and lipid metabolism(3, 5–7). Consequently, therapeutic strategies targeting obesity and related inflammatory and metabolic dysfunction are increasingly explored for OA management(3, 8–11). For instance, metformin, a widely used diabetes medication, has exhibited favorable effects on OA symptoms and progression in placebo-controlled trials of patients with knee OA and comorbid obesity(3, 10), as well as in a large-scale propensity score–matched cohort study involving patients with type 2 diabetes and OA(9). Similarly, a large, multicenter, placebo-controlled trial demonstrated that once-weekly subcutaneous semaglutide, a glucagon-like peptide-1 (GLP-1) receptor agonist, significantly reduced body weight and knee pain in patients with comorbid obesity and knee OA(8). Beyond direct receptor agonism, targeting upstream regulators such as the intestinal farnesoid X receptor (FXR) via administration of the secondary bile acid metabolite glycoursodeoxycholic acid (GUDCA) has been shown in preclinical models to promote GLP-1 secretion from intestinal L cells, facilitating its entry into the circulation and delivery to joints, thereby alleviating osteoarthritis(2).

Oligofructose and inulin are prebiotic fructans extensively studied for their ability to combat obesity and related metabolic abnormalities in both preclinical models and clinical settings(12–17). We recently conducted the first randomized placebo-controlled trial examining the effects of a 6-month oligofructose-enriched inulin intervention in adults with co-morbid obesity (BMI>30 kg/m^2^) and unilateral/bilateral knee OA (Kellgren-Lawrence grade II-III)(18, 19). Participants receiving prebiotic demonstrated significantly improved physical function, reduced trunk fat percentage, and altered gut microbiota and circulating metabolites(19). The microbial and metabolic mechanisms underlying these positive outcomes remain ill defined. We hypothesized that prebiotic supplementation enhanced phenotypic performance by modifying the composition and function of the gut microbiota in OA patients. To test this, we used a multi-omics approach to investigate the underlying mechanisms. We discovered that altered microbial carbohydrate utilization by the prebiotic responsive microbiome is linked to a systemic cytokine profile and distinct phenotypic changes via coordinated alterations in microbial and systemic metabolism. In a pre-clinical model of OA, we reveal a common inflammatory pathway is affected in humans and rats. Together, these mechanisms highlight the potential of utilizing the gut-joint axis for the treatment of knee OA(2, 19, 20).

## RESULTS

### Prebiotics reduced trunk fat percentage and improved physical performance

Given the significant impact of individual differences (baseline) on responses to microbiota-evoked stimuli or treatment(12, 21, 22), the present study included only the 30 participants (out of 54, 27 females and 3 males) who completed baseline and at least one of the two follow-up assessments and sample collections at months 3 and 6 from our previously reported randomized placebo-controlled trial(18, 19) (Fig. 1A, Table S1). We first examined how body composition and physical performance changed from baseline to 3 and 6 months between the prebiotic and placebo groups. While no significant changes in overall body composition including total body mass, body fat, lean mass, body fat percentage, and bone mineral density (BMD) were observed (Fig. 1B, Table S1), significant differences were observed in changes of trunk fat percentage at month 3 (*p*_Δmonth3_ = 0.044) and bone mineral content (BMC) at month 6 (*p*_Δmonth6_ = 0.047). Regarding physical performance, hand grip strength (*p*_Δmonth3_*=*0.015; *p*_Δmonth6_ = 0.008) and the Timed-Up-and-Go test (*p*_Δmonth3_ *<* 0.001; *p*_Δmonth6_ *<* 0.001) showed significant improvements in the prebiotic group compared to the placebo group throughout the intervention. The 40-m fast-pace walk test revealed a significant improvement in the prebiotic group at month 3 (*p*_Δmonth3_ *=* 0.020) (Fig. 1C, Table S1). No significant time effect was detected in the groups (Table S1). These favorable shifts in both obesity- and OA-related phenotypes following prebiotic intervention prompted us to investigate the underlying mechanisms further.

**Figure 1.**
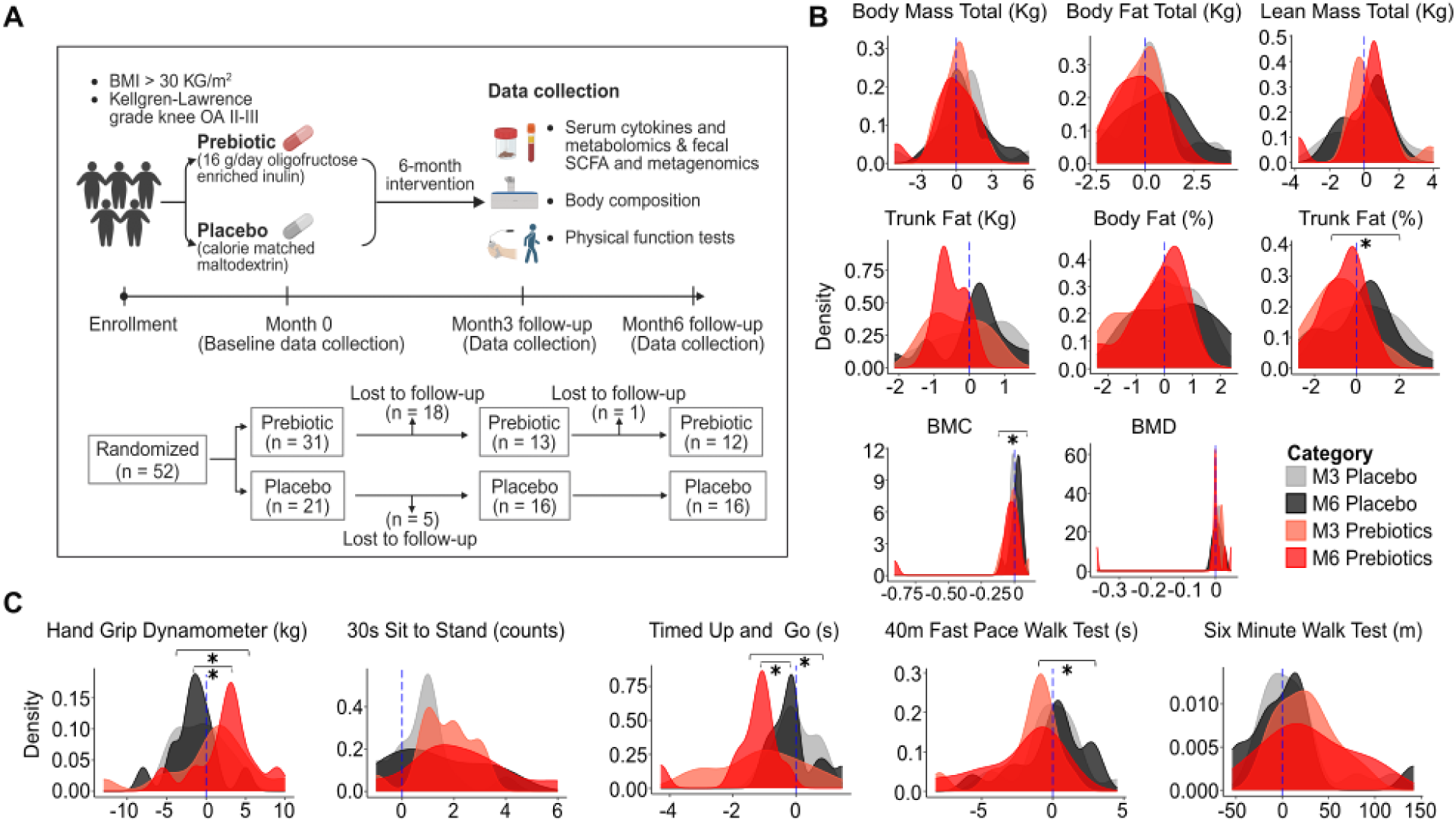
Overview of the 6-month prebiotic intervention and primary outcomes. **(A)** Timeline of the 6-month prebiotic intervention from enrollment to the 6-month follow-up. BMI, body mass index; OA, osteoarthritis; SCFA, short chain fatty acid. Changes in **(B)** body composition and **(C)** physical function at months 3 and 6 compared to baseline. Density plots indicate distribution of changes from baseline. Data with asterisk (*) represents a significant difference (*p* < 0.05).

### Prebiotic intervention shapes composition and function of the gut microbiome

We started by examining the impact of prebiotic intervention on its direct target, the composition and function of the gut microbiome. Metagenomic taxonomic composition and metabolic function were profiled using MetaPhlAn4(23) and HUMAnN3(24), respectively. The metagenomic potential for complex carbohydrate utilization was characterized by predicting Polysaccharide Utilization Loci (PULs) at the CAZyme gene cluster (CGC) level using dbCAN3(25). MetaPhlAn4 (species-level) taxonomy revealed no significant difference in α-diversity (Shannon index, Fig. 2A, upper). In contrast, the HUMAnN3 pathway showed significantly higher α-diversity at month 3 for the prebiotic group (*p* = 0.035, Fig. 2A, upper). Similarly, the α-diversity of dbCAN3 PUL prediction increased at month 3 in the prebiotic group (*p_month 0vs.3_ =* 0.005, Fig. 2A, upper, Table S2), while the placebo group showed a decline after month 3 (*p_month 0vs.6_* = 0.024; *p_month 3vs.6_* = 0.055, Fig. 2A, upper, Table S2). Two-dimensional NMDS of the Bray-Curtis distance-based β-diversity, calculated from MetaPhlAn4 taxonomy, HUMAnN3 pathway, and dbCAN3 PUL prediction explained 77.42%, 81.85%, and 85.35% of the variance in community dissimilarities, respectively (Fig. 2A, middle). PERMANOVA analysis on Bray-Curtis distance matrix identified individual variation and prebiotic intervention as significant factors explaining community dissimilarity across these profiles (*p* < 0.05, Fig. 2A, middle-bottom, Table S3).

**Figure 2.**
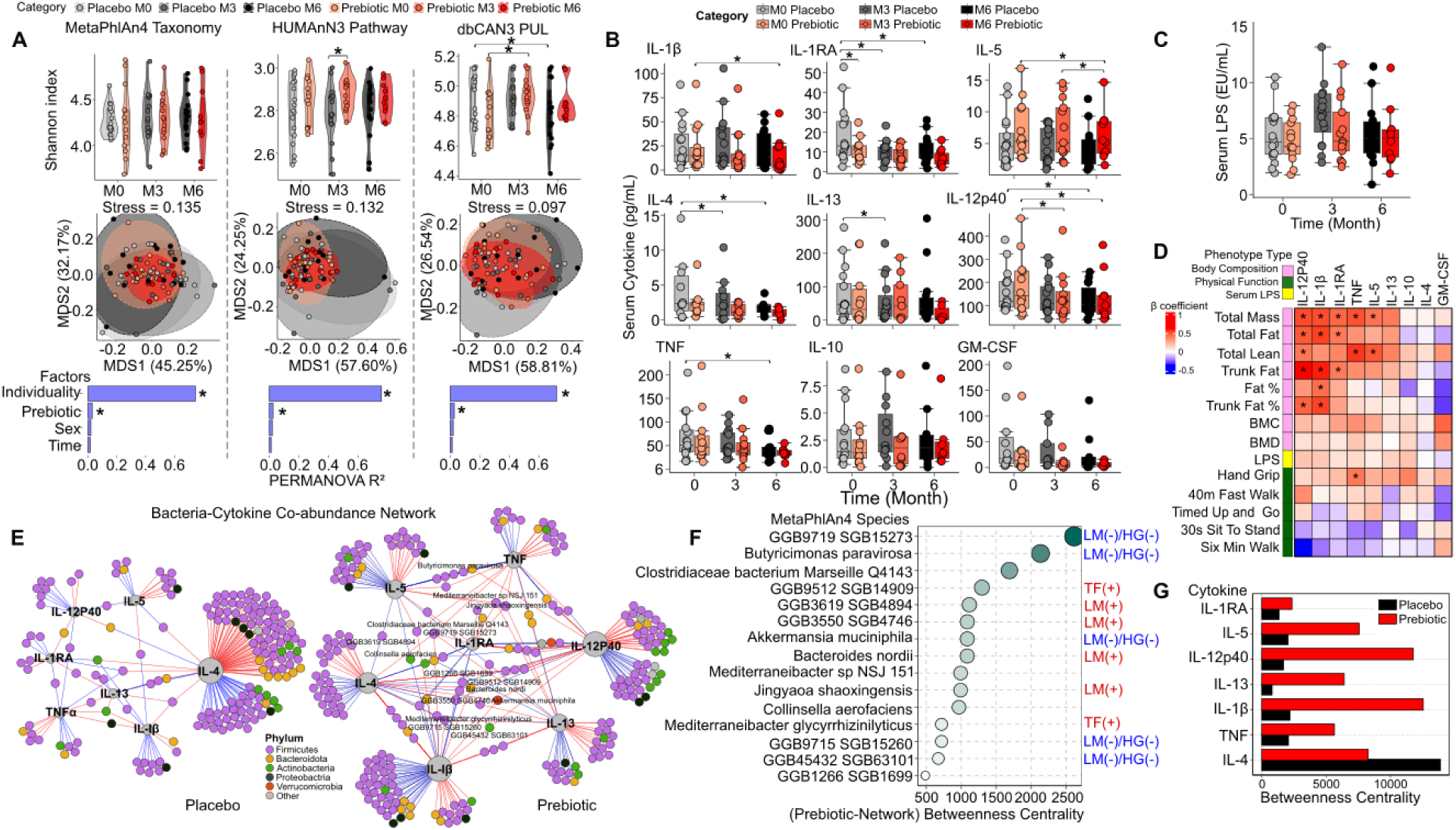
Impact of prebiotic intervention on fecal microbiome and systemic cytokine profile. **(A)** Compositional and functional changes in the fecal metagenome following prebiotic intervention. Violin plots (upper panels) illustrate changes in α-diversity (Shannon index) for metagenomic composition and function, presented as MetaPhlAn4 (species-level) taxonomy, HUMAnN3 metabolic pathway (unstratified), and dbCAN3 Polysaccharides Utilization Loci (PUL) prediction. Data with an asterisk (*) indicates significant differences (*p* < 0.05). Non-Metric Multidimensional Scaling (NMDS) plots (middle panels) depict inter-individual dissimilarities in Bray-Curtis distance matrix, calculated from MetaPhlAn4 taxonomy, HUMAnN3 metabolic pathway, and dbCAN3 PUL prediction, over time. Factorial analysis (lower panels) on Bray-Curtis distance matrix-based dissimilarities using PERMANOVA with 999 permutations. Factors with an asterisk (*) indicate a significant effect (*p* < 0.05). **(B)** Serum cytokines and **(C)** lipopolysaccharide (LPS) concentrations over time. Asterisks (*) indicate significant differences (*p* < 0.05), identified using mixed linear model. **(D)** Spearman’s correlations between serum cytokines, serum LPS and phenotypes. Data with asterisks (*) indicate significant correlations (*p* < 0.05). **(E)** Bacteria-cytokine co-abundance network of MetaPhlAn4 species and serum cytokine in placebo (left) and prebiotic (right) participants. Only significant Spearman’s correlations (*p* < 0.05) with coefficients > 0.45 are shown. Bacterial phylum is colored as per the legend. **(F)** Betweenness centrality of bacterial nodes in the bacteria-cytokine co-abundance network for prebiotic participants. LM, lean mass; HG, hand grip strength; TF, trunk fat %. The following (+)/(-) signs after phenotypes indicate species with significantly positive or negative (*p* < 0.05) Spearman’s correlations. **(G)** Betweenness centrality of cytokine nodes in the bacteria-cytokine co-abundance networks for prebiotic participants.

### Prebiotic responsive microbiome is linked to systemic cytokine profile and phenotypic changes

To move beyond microbiome-inferred direct impacts, we trained a phenotype-predictive random forest regression model with MetaPhlAn4 taxonomy, HUMAnN3 pathway, dbCAN3 PUL prediction, serum cytokines, and serum metabolites, individually or combined, as predictors. Combinations of serum cytokines with MetaPhlan4 taxonomy explained substantial variance in lean mass, trunk fat percentage, and hand grip strength (Fig. S1A, Table S4). Aligning with this, of the nine examined cytokines (Fig. 2B, Table S5) and lipopolysaccharides (LPS, Fig. 2C, Table S5) related to OA pathophysiology, seven cytokines showed divergent temporal changes between placebo and prebiotic groups. Notably, IL-1β decreased at month 6 in the prebiotic group, while its receptor antagonist IL-1RA decreased over time in the placebo group. IL-5 declined post-intervention in the prebiotic group, and IL-12p40 decreased in both groups. IL-4, IL-13, and TNF declined only in the placebo group. Changes in IL-1β, IL-1RA, and IL-12p40 correlated exclusively with body composition variables (excluding BMC/BMD), and hand grip strength (*p* < 0.05, Fig. 2D Cytokine-microbiome associations also differed between groups, with IL-4 dominating in the placebo and IL-1β in the prebiotic network (Fig. 2E, G). In the prebiotic network, 245 bacterial species correlated with cytokines (FDR < 0.05, R2 > 0.45, Fig. 2F), with 15 species changing post-intervention and 105 species significantly linked to phenotypic measures (Table S6-8). Phenotype-microbiome revealed that, of the 163 species significantly associated with hand grip strength, 99 also significantly associated with lean mass, while only 22 overlapped with trunk fat percentage (*p* < 0.05, Fig. S1C, Table S7-8). Most of the species positively associated with hand grip strength and lean mass were enriched in the prebiotic group, while negatively associated species decreased. In contrast, species linked to trunk fat percentage showed mixed patterns (Fig. S1B-C, Table S6-8). These results suggest that the prebiotic-responsive microbiome shares more associations with changes in lean mass and hand grip strength, with trunk fat percentage appearing more multi-faceted.

### Prebiotic intervention induces coordinated alterations in microbial and systemic metabolism

We next investigated phenotypic changes in association with metabolic alterations in the microbial community and host serum metabolome. Among four significantly altered phenotypes post-intervention, hand grip strength showed the strongest associations with HUMAnN3 pathways, followed by trunk fat percentage (FDR < 0.05, Fig 3A, Table S9). Specifically, microbial pathways involved in carbohydrate degradation, inorganic nutrient metabolism, and biosynthesis of carbohydrates, amino acids, secondary bile acids, cofactors, carriers, vitamins, and nucleosides and nucleotides were significantly associated with hand grip strength (FDR < 0.05, Fig 3A). Trunk fat percentage was mainly associated with degradation and biosynthesis of carbohydrate, and biosynthesis of secondary bile acids, cofactors, carriers, and vitamins (FDR < 0.05, Fig. 3A).

**Figure 3.**
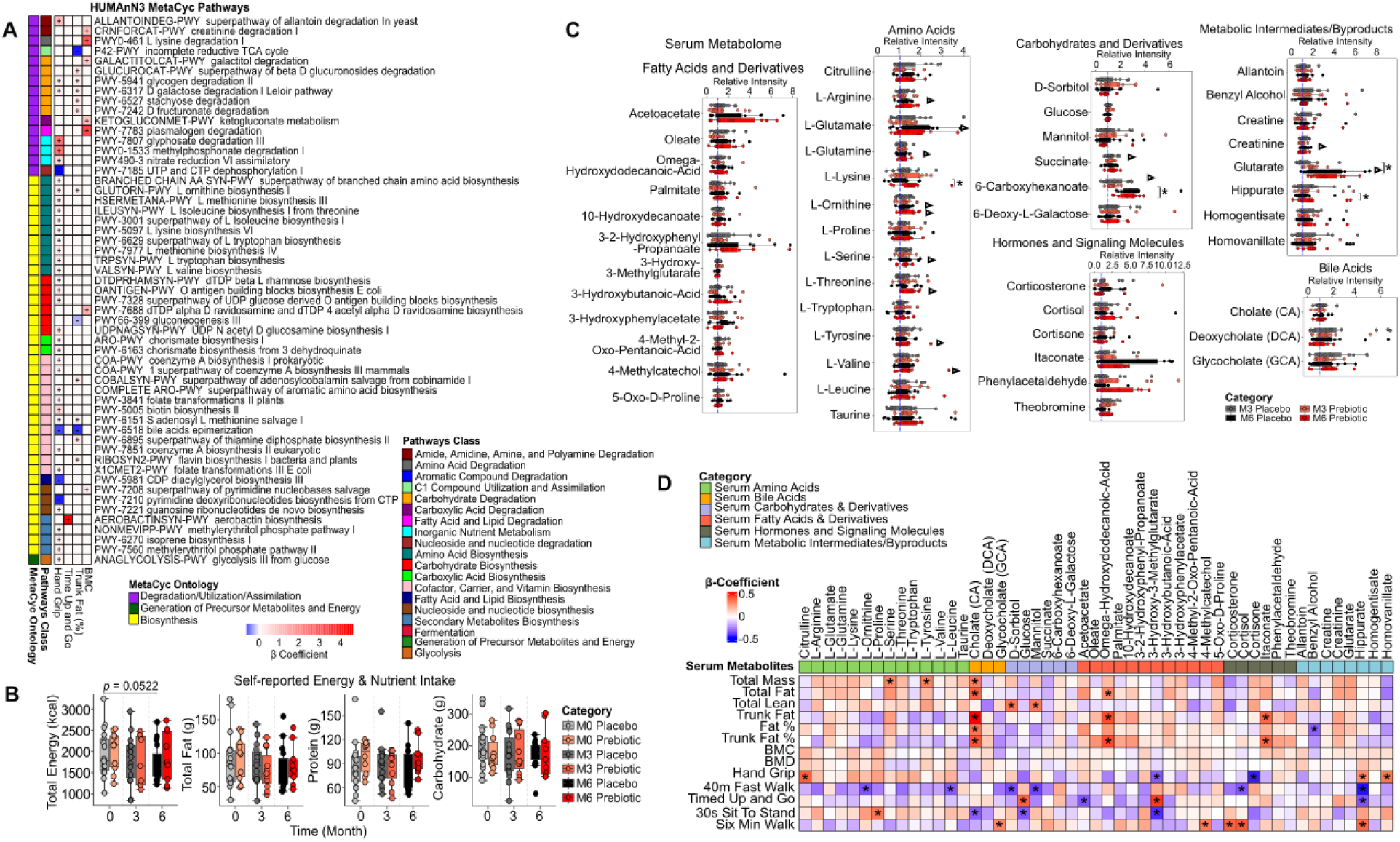
Phenotype-associated alterations in microbial metabolism and circulating metabolome. **(A)** Top HUMAnN3 pathways associated with prebiotic responsive phenotypes, identified using MaAsLin2. MetaCyc Ontology and Pathway Class are color-coded according to the key at the bottom-right. Heatmap column symbols indicate the significance of β coefficients: (+) for significantly positive (*p* < 0.05); (-) for significantly negative (*p* < 0.05). **(B)** Self-reported energy and nutrient intake following the intervention. **(C)** Serum metabolome changes following the intervention. Triangles (Δ) indicate significant differences compared to baseline (*p* < 0.05), and asterisks (*) indicate significant differences (*p* < 0.05) in general comparisons, identified using linear mixed model. **(D)** Spearman’s correlations between baseline-normalized serum metabolome and phenotypes, with asterisks (*) denoting significant correlations (*p* < 0.05). The category of metabolites is color-coded according to the key in the top-left.

Despite no significant differences in energy and macronutrient intake between groups (Fig. 3B), the serum metabolome differed between prebiotic and placebo groups, primarily in amino acids at month 6, carbohydrates and derivatives at month 3, and metabolic intermediates/byproducts at month 6 (Fig. 3C, Table S10). L-arginine, L-ornithine, L-glutamate, L-glutamine, and L-serine increased significantly in the placebo group, while L-ornithine, L-threonine, L-tyrosine, and L-valine increased significantly in the prebiotic group (*p* < 0.05, Fig. 3C). At month 6, L-lysine was significantly higher in the prebiotic group. Succinate and 6-carboxyhexanoate increased significantly in the placebo group at month 3, and 6-carboxyhexanoate was higher in the placebo group at month 6 (*p* < 0.05, Fig. 3C). At month 6, creatinine and glutarate increased significantly in the placebo group, and hippurate was higher in the prebiotic group (*p* < 0.05, Fig. 3C).

Correlations between baseline-normalized serum metabolome and phenotypic changes revealed significant associations between changes in body mass and fat, particularly trunk fat (*p* < 0.05) and the primary bile acid cholate (CA), fatty acid derivatives omega-hydroxydodecanoic acid and 3-hydroxy-3-methylglutarate (HMG), and the immunomodulatory metabolite itaconate(26–28) (Fig. 3D). Changes in L-serine and L-tyrosine positively correlated with total body mass changes, while D-sorbitol and mannitol changes positively correlated with lean mass changes. Benzyl alcohol was the only metabolite negatively correlated with body fat percentage (*p* < 0.05, Fig. 3D). Changes in citrulline, L-ornithine, L-proline, L-leucine, glycocholate (GCA), D-sorbitol, mannitol, 4-methylcatechol, corticosterone, cortisol, acetoacetate, hippurate, and homovanilate were positively correlated with improvements in physical function, whereas changes in CA, glucose, HMG, and cortisone were negatively correlated with improvements in physical function (*p* < 0.05, Fig. 3D). The coordinated associations between phenotypic changes and microbial/systemic metabolism suggest a central factor that shapes the co-metabolism between host and microbial community.

### Prebiotics alter microbial carbohydrate utilization, shifting systemic metabolic and phenotypic outcomes

To explore how prebiotics shifted host and microbial co-metabolism, we reconstructed the longitudinal changes in the metagenomic machinery responsible for utilizing complex carbohydrates at the CAZyme gene cluster (CGC) level. By querying sample-wide CGCs against the dbCAN-PUL database, we identified 369 PULs, with 14 containing experimentally characterized glycan substrates that showed significant group or temporal differences (Fig. 4A, Table S11). Among these, two β-glucan degrading PULs (PULs 0019 and 0585) showed significant differences (*p* < 0.05) between the prebiotic and placebo groups at month 3. In the placebo group, significant time effects were identified in PULs degrading α-glucan (PULs 0371, 0633, 0638, and 0639), galactan (PULs 0323 and 0651), and host glycan (PULs 0196 and 0366). In contrast, for the prebiotic group, significant time differences were found in PULs degrading β-mannan (PUL0179), xylan (PULs 0392 and 0619), and pectin (PUL0189) (Fig. 4A, Table S11).

**Figure 4.**
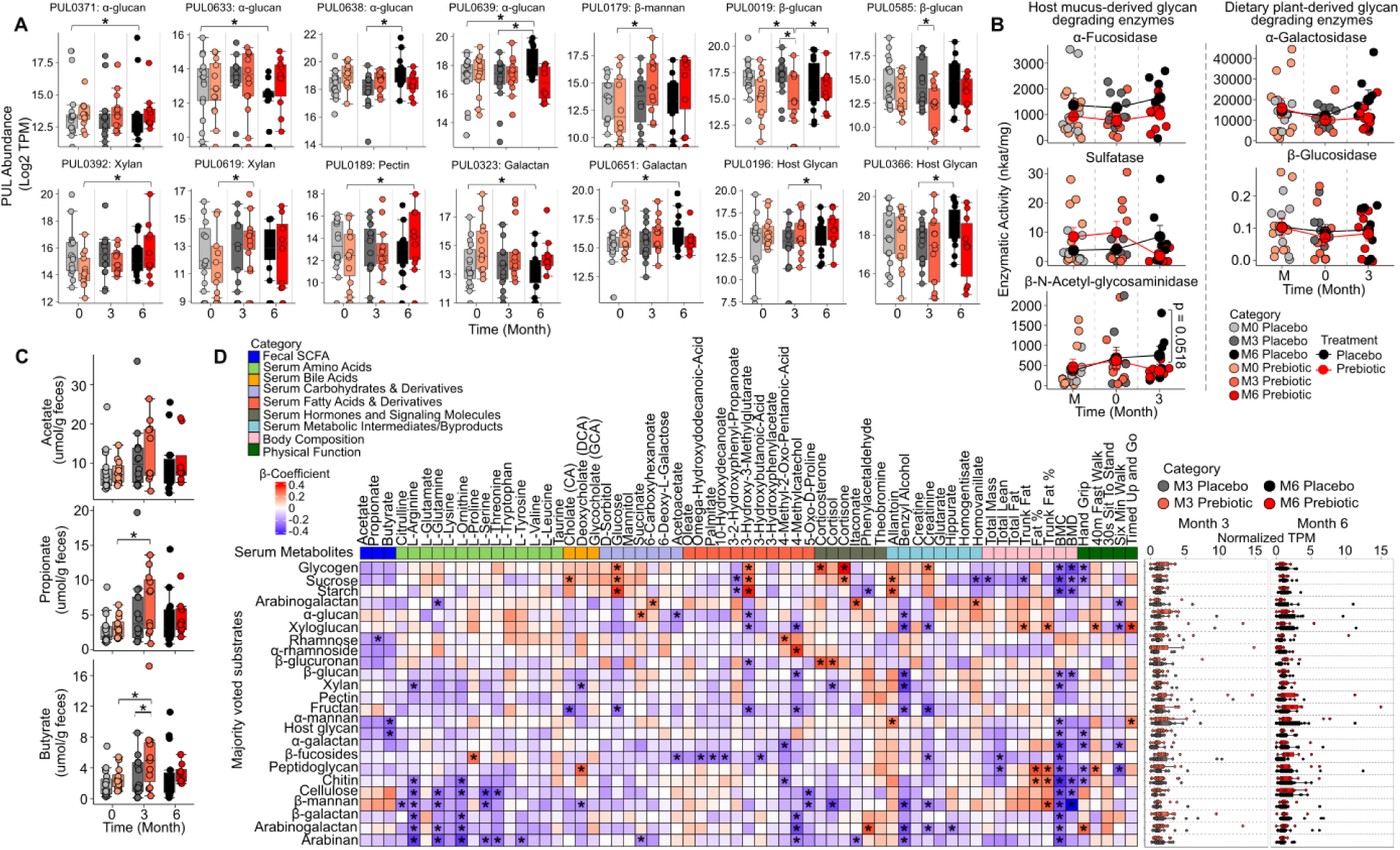
Microbial glycan-degrading activity profiling and relation to circulating metabolism and phenotypes. **(A)** Polysaccharides Utilization Loci (PULs) with significant differences (FDR < 0.05) between prebiotic and placebo participants over time. Data was represented as Log_2_TPM, with asterisks (*) denoting significant differences (*p* < 0.05). **(B)** Activities of microbial host mucus- and dietary plant-derived glycan degrading enzymes measured using 4-nitrophenyl-coupled substrates. Jitter dots represent individual measurements, and the line connected dots show the mean enzymatic activity for placebo (black) and prebiotic (red) groups at each time point. **(C)** Fecal short-chain fatty acid (SCFA) concentrations over time, with asterisks (*) denoting significant differences (*p* < 0.05). **(D)** Spearman’s correlations between baseline-normalized CAZyme subfamilies’ majority voted substrates at the CGC level and fecal SCFA, serum metabolites, and phenotypes, with asterisks (*) denoting significant correlations (*p* < 0.05) (left panel). The right panel demonstrates the baseline-normalized abundance change of each majority-voted substrates over time.

To validate the divergence in carbohydrate utilization predictions, we measured the activity of three mucus-glycan degrading bacterial enzymes (α-fucosidase, sulfatase, and β-N-acetyl-glycosaminidase), along with two plant-derived glycan degrading enzymes, α-galactosidase and β-glucosidase following the intervention (Fig. 4B, Table S12). While no significant changes were observed in other enzymatic activities, β-N-acetylglucosaminidase, which hydrolyzes the β-1,4 or β-1,3 bonds between N-acetylglucosamine residues in glycans and glycosaminoglycans showed a trend of lower activity in the prebiotic group at month 6 (*p* = 0.0518, Fig. 4B, Table S12). Direct quantification of fecal SCFA showed that propionate and butyrate concentrations increased significantly in the prebiotic group at month 3 compared to baseline (*p* < 0.05, Fig. 4C, Table S5).

In addition to PULs predictions, we quantified sample-wide carbohydrate feedstock utilization by summing the abundance of CGCs with the same predicted substrate, applying a majority voting approach across all CAZyme subfamilies per CGC, and examined their associations with serum metabolome and participant phenotypes (Fig. 4D). Overall, participants’ microbiome exhibited the greatest metagenomic potential for degrading host glycans, followed by xylene, β-glucan, pectin and starch (Fig. S2). Shifts in carbohydrate utilization were broadly correlated with changes in fecal SCFA concentration, serum metabolome, and phenotypic measures. Importantly, changes in α-mannan and host glycan utilization negatively correlated with fecal butyrate concentration, and/or performance on the timed-up-and-go test and hand grip strength (*p* < 0.05, Fig. 4D). Shifts in microbial utilization of chitin, cellulose, β-mannan, β-galactan, arabinogalactans and arabinan were found most often to negatively correlate with serum L-arginine, L-glutamine, L-ornithine, L-serine, and L-threonine (*p* < 0.05, Fig. 4D). Arabinogalactans also negatively correlated with 4-methylcatechol, benzyl alcohol, and creatinine and positively correlated with itaconate and hand grip strength (*p* < 0.05, Fig. 4D). Changes in utilization of glycogen, sucrose, and starch positively correlated with serum glucose and HMG (3-hydroxy-3-methylglutarate), coupled with cholate (CA), cortisone, or allantoin, and negatively correlated with BMC, BMD, and/or hand grip strength (*p* < 0.05, Fig. 4D). Notably, changes in microbial utilization of fructan (prebiotic) were the only carbohydrates negatively correlated with changes in serum CA, glucose, HMG, and creatinine (*p* < 0.05, Fig. 4D). These findings underscore the pivotal role of the shifted complex carbohydrates utilization following prebiotic intervention in shaping microbial and systemic metabolism, particularly with respect to bile acids, SCFAs, and AAs, ultimately leading to phenotypic improvements.

### Metagenomic reconstruction couples fructan degradation with bile acid metabolism, butyrate concentration, and host-glycan degradation

We reconstructed 596 high-quality metagenomic-assembled genomes (MAGs) to explore how prebiotic-driven fructan degradation was intrinsically linked to altered bile acid metabolism, butyrate concentration, and host glycan utilization (Fig. 5A, Table S13). We identified 57 fructan-degrading MAGs primarily from *Bacteroides*, *Blautia*, *Bifidobacterium*, *Anaerostipes*, and *Lacticaseibacillus*, and 291 host-glycan-degrading MAGs dominated by *Bacteroides*, *Eisenbergiella*, *Alistipes*, *Enterococcus*, *Butyricimonas*, *Gemmiger*, *Collinsella*, and *Akkermansia* (Fig. 5A). Among the fructan-degraders, *B. uniformic* was the dominant species involved in bile acid epimerization via its possession of the 7α-hydroxysteroid dehydrogenase (7αHSDH) gene, which was significantly enriched in the prebiotic group (*p* < 0.05, Fig. 5B, Table S14).

**Figure 5.**
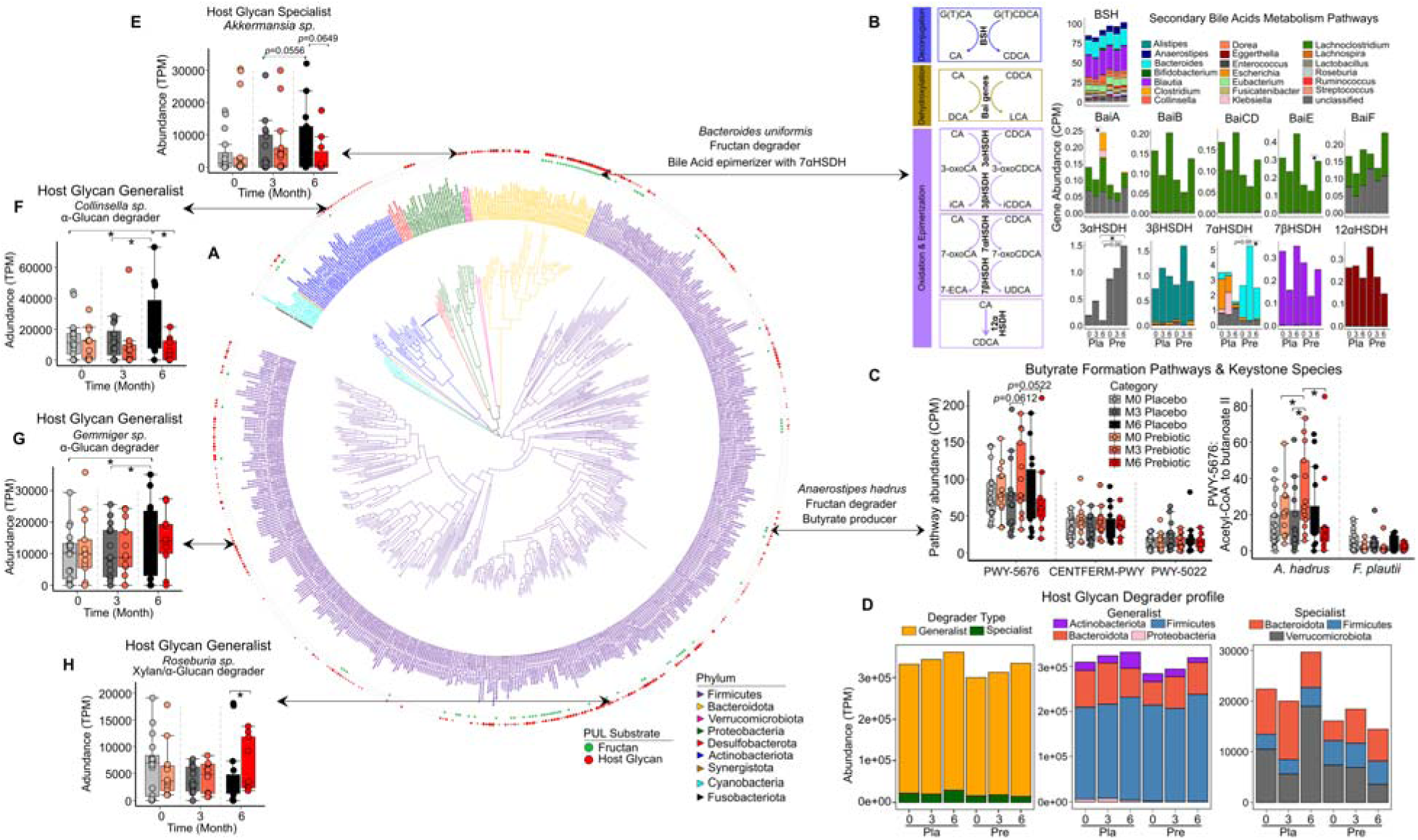
Metagenomic reconstruction of carbohydrates and secondary bile acid metabolism. **(A)** Phylogeny and predicted potential for fructans and host glycan utilization of metagenomic assembled genomes (MAG) recovered from participant stool samples. The phylogenetic tree and the taxonomic assignment of the 596 reconstructed high-quality MAGs (completeness ≥ 70%, contamination < 5%) are shown as the two inner layers, color-coded by phyla as indicated by the legend in the lower right. The outer two layers depict the ranking for fructan (green) and host glycan (red) degradation potentials based on the number of fructan- and host glycan- degrading PULs in each MAG. Major pathways and bacterial species involved in (**B)** secondary bile acid metabolism and **(C)** butyrate formation, with asterisks (*) denoting significant correlations (*p* < 0.05) determined by mixed linear model. **(D)** Longitudinal changes of host glycan degrader type (left) and specific changes of each type at phylum level (middle and right), with color coding indicating corresponding type or phylum. Representative genera of host glycan specialist **(E)** and generalist **(F-H)** with significant differences following intervention, with asterisks (*) denoting significant correlations (*p* < 0.05) determined by mixed linear model.

*Anaerostipes hadrus*, another fructan-degrader, was significantly enriched in the prebiotic group at month 3 (*p* < 0.05, Fig. 5C), and was the primary contributor to butyrate production via the acetyl-CoA fermentation to butanoate II pathway (PWY-5676). Among the 291 host glycan MAGs, 12 were classified as host glycan specialists(29), carrying host-glycan PULs as the primary carbohydrate utilization machinery (comprising >67% total PULs), while the remaining 279 were host glycan generalists (Fig. 5D, Table S13)(29–33). The specialist group, mainly from *Verrucomicrobiota*, *Bacteroidota*, and *Firmicutes* (*Bacillota*), showed no significant differences (Fig. 5D), while the generalist consortium, dominated by *Firmicutes*, *Bacteroidota*, *Actinobacteriota* (*Actinomycetota*), and *Proteobacteria* (*Pseudomonadota*), showed significant temporal differences between the groups. *Firmicutes* generalists increased in the prebiotic group from month 3 to 6, whereas *Actinobacteriota* generalists increased in the placebo group over time (Fig. 5D). At the genus level, specialist genus *Akkermansia* showed an increasing trend in the placebo group from month 3 to 6 (*p* =0.0556) and a higher abundance in the placebo group at month 6 (*p* =0.0649, Fig. 5D-E). In contrast, *Roseburia*, a top-abundant generalist and xylan and α-glucan degrader, significantly increased in the prebiotic group at month 6 (*p* < 0.05, Fig. 5H). *Collinsella, and Gemmiger,* other top-abundant generalists and α-glucan degraders, significantly increased in the placebo group over time (*p* < 0.05, Fig. 5F-G). These findings provide compelling metagenomic evidence that the divergent shifts in microbial degradation of dietary-versus host-derived glycans between the placebo and prebiotic groups are directly coupled with the observed differences in serum bile acids and fecal butyrate.

### Multi-omics factor analysis identifies key signatures driving phenotypic improvements

Next, we integrated our multiomic datasets using unsupervised Multi-Omics Factor Analysis (MOFA2) to fully exploit inter-participant variability. The MOFA2 model captured two latent factors, revealing that shifts in microbial diet- versus host-derived glycan degradation and L-arginine/L-ornithine metabolism played central roles in linking prebiotic intervention to phenotypic outcomes. Latent factor 1 explained 40.3% of the variance in prebiotic and 29.5% in placebo participants (Fig. 6A). It captured reduced host-glycan degradation and increased microbial synthesis of L-arginine and L-ornithine, two amino acids central to citrulline metabolism (Fig. 6C, upper), and correlated with improved physical performance in the hand grip strength and the 30-second sit-to-stand tests (Fig. 6B, left). Latent factor 2 explained 41.3% of the variance in prebiotic and 6.6% in placebo participants (Fig. 6A). It reflected SCFA-mediated shifts that mitigated excessive host-glycan foraging, dampened IL-1β-driven inflammation, and shifted microbial metabolic adaptations favoring cellular survival and stress responses (Fig. 6C, bottom). These key signatures align closely with our knowledge-based findings, offering a comprehensive understanding of the mechanisms underlying the phenotypic improvements post-intervention.

**Figure 6.**
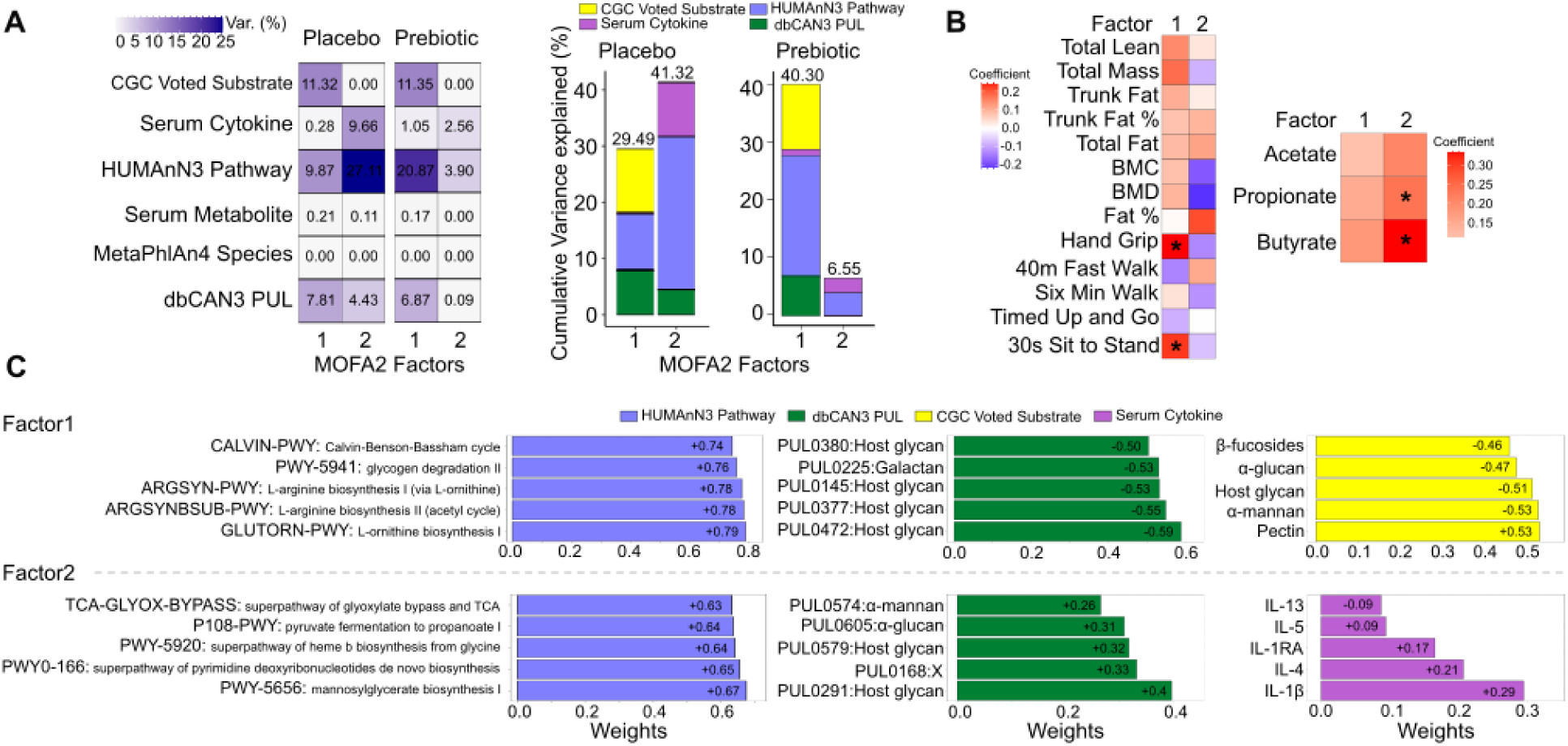
Unsupervised multi-omics integration using Multi-Omics Factor Analysis (MOFA2) coupled with group- and time-aware MEFISTO. **(A)** Latent factors captured using MOFA2 coupled with MEFISTO group- and time-aware settings, across different datasets from prebiotic and placebo participants following the intervention. **(B)** Correlation of latent factors with fecal SCFA and phenotypic measures, with color coding reflecting co-efficient values. **(C)** Features with top weights correlating with latent factors, with the color key indicating the view type. Numbers representing weights values and directions (positive +, negative -).

### Prebiotics protect knee joint health in a diet-induced obesity rat model

Lastly, we aligned our findings in humans with results from a previous animal study examining the protective effect of prebiotics on knee joint health in a diet-induced obesity rat model(34) (Fig. 7A). After a 12-week intervention, rats fed a high-fat/high sucrose diet (HFS) showed significant increases in serum IL-1β and IL-10 (*p* < 0.05, Fig. 7B, Table S15), as well as higher TNF and IL-5 levels compared to rats fed HFS with 10% wt/wt oligofructose prebiotic (HFS+P, *p* < 0.05, Fig. 7B, Table S15). Synovial fluid IL-1β trended higher in HFS rats (*p* = 0.0860, Fig. 7C). While total bacterial abundance remained unchanged, HFS+P significantly decreased abundance of the host-glycan specialist *A. muciniphila* but increased abundance of the generalist *Roseburia* spp., resulting in a lowering of the *A. muciniphila/Roseburia* spp. ratio (*p* < 0.05, Fig. 7D-E). Joint histology revealed no significant difference in total joint histologic score and bone changes (Table S11), but reduced lateral tibial plateau cartilage degeneration (*p* < 0.05, TableS16, Fig. 7F) and less synovium membrane thickening (*p* < 0.05, TableS16, Fig. 7G) in HFS+P rats. The *A. muciniphila/Roseburia* spp. ratio correlated significantly with serum IL-1β, IL-10, IL-4, and IL-13 (*p* < 0.05, Fig. 7H, Table S15) and synovium thickening scores (*p* < 0.05, Fig. 7I, bottom). Additionally, *A. muciniphila* abundance correlated significantly with lateral tibial plateau cartilage scores (*p* < 0.05, Fig. 7I, top). These pre-clinical findings in an obese rat model suggest a protective effect of prebiotic intervention against OA onset and progression through a common inflammatory pathway we observed in humans.

**Figure 7.**
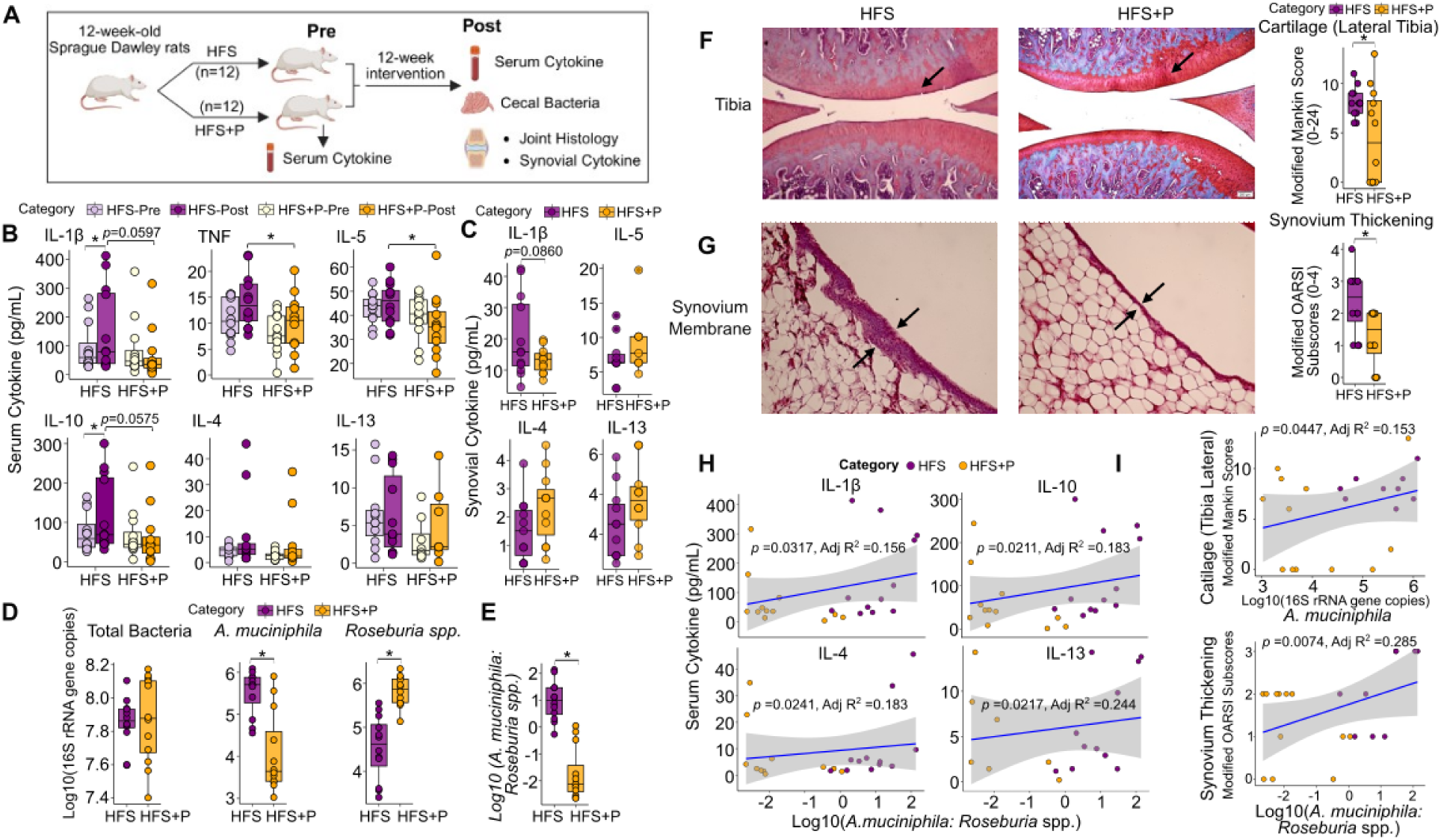
Protective effects of prebiotic consumption on knee health in a diet-induced obesity rat model. **(A)** Schematic overview of the rat study design; HFS, high fat/high sucrose diet; HFS+P, HFS+10% (wt/wt, Orafti P95 oligofructose) prebiotic; Pre/Post, Pre-/Post-intervention. **(B)** Serum pre- and post-intervention and **(C)** synovial post-intervention cytokine concentrations. Asterisks (*) indicate significant differences (*p* < 0.05), identified using (mixed) linear model. **(D)** Abundance of total cecal bacteria, *Akkermansia muciniphila*, and *Roseburia* spp., expressed as Log10 (16S rRNA gene copes per 20 ng DNA). **(D)** The *A. muciniphila/Roseburia* spp. ratio, asterisks (*) indicate significant differences (*p* < 0.05), identified using the Kruskal-Wallis test. Representative histological images (left) and quantification scores (right) of knee joints reflecting **(F)** tibial lateral cartilage degradation and **(G)** synovial membrane thickening. **(H)** Regressions between the *A. muciniphila*/*Roseburia* spp. ratio and serum cytokines, only regressions with *p* < 0.05 were shown. **(I)** Regressions between *A. muciniphila* abundance and Modified Mankin score for tibial lateral cartilage (upper panel) and between the *A. muciniphila*/*Roseburia* spp. ratio and Modified OARSI subscores for synovium membrane changes (bottom panel).

## DISCUSSION

Modulating inflammatory pathways(35–40), reducing enzymatic cartilage degradation(37, 38), and stimulating regeneration of extracellular matrix components such as proteoglycan, collagen and hyaluronic acid(39, 40) remain primary targets in developing disease-modifying OA drugs. However, no pharmacological agents targeting these mechanisms have yet received regulatory approval. Obesity-related OA now accounts for a substantial proportion of OA cases and is associated with accelerated disease progression and earlier need for total knee replacement(41–43). Effective treatment for obesity-related OA necessitates long-term follow-up to monitor disease progression and potential side effects³¹, imposing significant financial, logistical, and patient adherence challenges on healthcare systems. In this context, our randomized controlled trial^19^ demonstrated the potential of a 6-month prebiotic intervention as a conservative, easy to adhere to management strategy for adults with comorbid knee OA and obesity. Here we show that daily intake of 16 g of oligofructose-enriched inulin increased diet-derived carbohydrates available to the gut microbiome, mitigated excessive host-glycan degradation and mucosal barrier disruption, and reduced systemic inflammation and metabolic dysregulation, ultimately improving metabolic health and physical function. Notably, the protective effect of prebiotic against OA onset and progression was further supported in a diet-induced obesity rat model, as evidenced by the amelioration of tibia cartilage degeneration and synovium membrane thickening.

Mechanistically, β-(2→1) glycosidic bonds in inulin-type fructans resist digestion in the small intestine, allowing most prebiotic fibers to reach the large intestine, where they are degraded by microbial fructan-degrading PULs with GH32 β-fructanases(44). Reconstruction of the metagenomic machinery responsible for complex carbohydrate utilization revealed that total fructan degradation was negatively correlated with serum levels of cholate (a primary bile acid), glucose, and 3-hydroxy-3-methylglutarate (HMG), metabolites that correlated unfavorably with body fat and physical performance. The inverse relationship between microbial fructan degradation and systemic cholate was primarily driven by *B. uniformis*, a fructan-degrader and dominant species that epimerizes primary bile acids through 7α-hydroxysteroid dehydrogenase (7αHSDH). These observations align with previous reports showing that fructan supplementation reduced weight gain and improved glucose metabolism in mice by promoting bile acid epimerization, which activates the TGR5-GLP1 (Takeda G protein-coupled receptor 5 receptor-glucagon-like peptide-1) signaling axis(45). The relationship between bile acid metabolism and GLP-1 is further strengthened by observations from two human cohorts showing altered microbial bile acid metabolism with reduced glycoursodeoxycholic acid (GUDCA) in osteoarthritis and lower risk of joint replacement in patients supplemented with ursodeoxycholic acid (an FDA approved drug and precursor of GUDCA)(2). In mice, farnesoid X receptor (FXR) inhibition and/or intra-articular injection of the GLP-1 receptor agonist liraglutide, mitigates cartilage degradation(2). Based on these results, there is growing interest in the therapeutic potential of targeting microbial bile acid metabolism, either by supplementation with the secondary bile acid GUDCA or with *C. bolteae*, a bacteria responsible for production of ursodeoxycholic acid (GUDCA precursor) in the human gut. Important to our findings is that prebiotic fructans are known to increase intestinal L cell numbers, enhance GLP-1 secretion, and alter bile acid metabolism(46–49), potentially explaining one pathway through which the beneficial effects of oligofructose-enriched inulin were manifest in our participants.

Another key mechanistic mediator of the prebiotic effects was through the enrichment of *A. hadrus*(12), a fructan-degrader and primary butyrate producer via acetyl-CoA fermentation(12, 50, 51). Fecal butyrate concentrations negatively correlated with host-glycan degradation, which diverged between placebo and prebiotic participants, as indicated by PUL predictions and glycan-degrading enzyme activities. Within the host-glycan degrading consortium, generalists such as *Bacteroides* spp.(33, 52), *Roseburia* spp(30), and *A. hadrus*(32, 53), thrive on diverse carbohydrates, including fructan, α-glucan, xylan, β-mannan, pectin, and β-glucan, while specialists such as *A. muciniphila*(29, 54, 55), and *Barnesiella intestinihominis*(29), expand primarily by foraging mucin when diet-derived carbohydrates are limited. While *A. muciniphila* is a dominant mucolytic commensal species under homeostasis(54, 56), its excessive expansion renders host susceptibility to mucosal pathogen penetration(29), inflammation(57), and food allergy(58) in the context of epithelial barrier damage. In our study, prebiotic fiber supplementation protected participants with comorbid knee OA and obesity from excessive host-glycan degradation, ameliorating intestinal barrier dysfunction and systemic inflammation, as evidenced by higher serum hippurate levels, a metabolic and intestinal health marker(59, 60) and reduced circulating IL-1β, a major OA-related pro-inflammatory cytokine(61, 62).

Coupled with altered carbohydrate metabolism, amino acid metabolism dynamics, particularly biosynthesis of L-arginine, L-ornithine, and citrulline, in both circulation and gut microbiota were modulated by prebiotic intervention, correlating with improved physical function. Circulating citrulline, synthesized mainly from L-ornithine and carbamoyl phosphate(63), positively correlated with hand grip strength improvements, likely attributed to its suppressive role in proinflammatory macrophage activation(63–65). Upon pathogen invasion, macrophages convert L-arginine to citrulline via inducible nitric oxide synthase (iNOS), concurrently producing nitric oxide (NO), a key antimicrobial effector(65). Notably, reduced serum L-arginine was present in placebo participants at month 6, reflecting a potentially heightened macrophage activation in this group. This aligns with the protective effects of prebiotic fiber against excessive host-glycan degradation and mucosal bacterial penetration, thereby mitigating IL-1β-driven inflammation.

The unsupervised MOFA2 model placed shifts in diet- versus host-derived glycan degradation and reduced pro-inflammatory immune responses, mainly driven by IL-1β, as the central mechanism linking prebiotic intervention to phenotypic improvements. IL-1β is a key proinflammatory cytokine involved in OA pathophysiology, particularly articular cartilage matrix degeneration(66, 67). This is consistent with a previous animal study showing the protective effect of prebiotic fiber on knee health in a diet-induced obesity rat model(34). Upon obesity induction with a high fat/sucrose diet, prebiotic supplementation dampened serum and synovial IL-1β increases, and other pro-inflammatory cytokines, modulated the microbiota by mitigating the increase in host-glycan specialist *A. muciniphila* and enriching generalist *Roseburia* spp., thereby lowering the *A. muciniphila/Roseburia* spp. ratio. Importantly, this was significantly associated with reduced lateral tibial plateau cartilage degeneration and less synovial membrane thickening. The *A. muciniphila/ Roseburia* spp. ratio and *A. muciniphila* abundance correlated significantly with serum cytokines, including IL-1β, and synovial thickening and cartilage degeneration scores, highlighting a mechanistic link between microbial changes, immune modulation, and joint tissue protection.

## CONCLUSION

Our findings highlight the promising therapeutic potential of prebiotic supplementation as both a conservative management strategy that improves physical function and metabolic health in adults and a preventive approach for obesity-related knee OA by protecting against OA onset and progression in a preclinical model. Prebiotic-induced shifts in microbial carbohydrate utilization were linked to improved physical and metabolic outcomes. With the rising prevalence of obesity-related OA and the lack of effective treatment options, our study provides important mechanistic insight into prebiotic-associated improvements occurring along the gut-joint axis.

## METHODS

### Participants

A total of fifty four adults (49 females, 5 males) aged 30-75 with a BMI > 30 kg/m² and a diagnosis of unilateral or bilateral knee osteoarthritis (grades II-III) according to the Kellgren-Lawrence radiographic rating scale were recruited from Calgary, Alberta, Canada and surrounding area between 2018 and 2020. The exclusion criteria included injury-associated knee OA, previous surgeries (knee surgery, bariatric surgery, or other gastrointestinal surgeries), active use of weight loss medication, use of prebiotics, probiotics or antibiotics in the previous 3 months, active infection or malignancy, pregnancy, and lactation. The full details of this double-blind, placebo-controlled trial have been previously published(18, 19). Computer generated numbers were used to randomize eligible participants to either prebiotic (16 g/day Orafti Synergy1 oligofructose-enriched inulin, Beneo, Mannheim, Germany) or a calorie-matched placebo (6.6 g/day Agenamalt 20.222 maltodextrin, Agrana Starch, Konstanz, Germany) for 6 months. The powders were mixed in water and consumed daily. Analysis for the present study was conducted on a subset of participants (n = 30) who completed the baseline assessment and at least one of the two follow-up physical function measurements and sample collections at months 3 and 6 (18, 19). The sample size of the trial was determined to maintain statistical power of 0.80 and a probability of 0.05, based on previously reported data regarding anticipated changes in body fat with prebiotic fiber supplementation in adults with overweight or obesity(68). Ethical approval was provided by the Conjoint Health Research Ethics Board of the University of Calgary and was prospectively registered at ClinicalTrial.gov (NCT04172688).

### Study design

Changes in physical function were assessed through participant’s performance on a subset of the Osteoarthritis Research Society International (OARSI) tests(69) at baseline, month 3, and month 6. Physical function tests included the 30s chair stand test for lower body strength, the 40m fast-paced walk test for short walking distance, the timed up & go test for ambulatory activity (assessing strength, agility, and dynamic balance), the hand grip strength as a measure of overall body muscle strength and physical function, and the 6-min walk test for aerobic capacity and long distance walking activity. On the same test days, serum samples were collected to profile circulating inflammatory and metabolomic markers in relation to prebiotic intervention induced changes in metabolism and physical functions. Stool samples were collected at home using stool collection kits and brought to the laboratory on test days followed by storage at -80.

### Physical function tests and participant reported outcomes

Each of the five performance-based tests (30s chair stand test, the 40m fast-paced walk test, the timed up & go test, the hand grip strength, and the 6-min walk test) were completed at each visit (months 0, 3, and 6). Full details of the tests have been previously published^1^. In addition to physical function tests, participants were asked to report on the following outcomes: (1) knee pain intensity, rated on a scale from 0 (no pain) to 10 (worst possible pain), according to the Numerical Pain Rating Scale (NPRS) for knee pain; (2) Abdominal discomfort, bloating, flatulence, stomach rumbling, and frequency of bowel movements, using a standardized gastrointestinal symptoms questionnaire(70) (3) Quality of life, assessed through the 36-Item Short Form Health Survey (SF-36)(71), a self-administered tool measuring health across multiple dimensions, including functional status, well-being, and overall health evaluation; (4) Dietary intake, with participants maintaining their usual diet and recording their food consumption using a 3-day food record at baseline, 3, and 6 months; (5) Physical activity, with participants reporting their average weekly exercise intensity (strenuous, moderate, mild) over the past month, using a modified version of Godin’s Leisure-Time Exercise Questionnaire(72).

### Serum metabolome

A Q Exactive™ HF Hybrid Quadrupole-Orbitrap™ Mass Spectrometer (Thermo-Fisher, Waltham, MA USA) coupled to a Vanquish™ UHPLC System (Thermo-Fisher) was used to perform serum metabolomics at the Calgary Metabolomics Research Facility (Calgary, Canada), as previously described(73). Briefly, the injection volume of samples was 2 uL. Chromatographical separation of metabolites was performed on a Syncronis HILIC UHPLC column (2.1mm x 100 mm x 1.7 um, Thermo-Fisher) at a flow rate of 600 uL/min using a binary solvent system. Data were acquired in both positive and negative full scan mode at a resolution of 240,000, scanning from 50-750m/z. Spectrum data was analyzed by El-MAVEN software package (74). Metabolite peak assignments were determined by matching observed m/z signals (+/-10ppm) and chromatographic retention times of authentic standards (Sigma-Aldrich) with observed metabolite signals. Metabolites with intensity lower than three times intensity of Mid-Blanks were discarded. The absolute concentration of retained metabolites was calculated by fitting with an eight-point quantification curve of authentic standards.

### Serum Lipopolysaccharides and inflammatory markers

Serum lipopolysaccharides (LPS) levels were measured using the Pyro-Gene recombinant factor C endotoxin detection kit (Lonza, Walkersville, MD) following the manufacturer’s protocol. Serum inflammatory biomarkers were profiled using the 42-plex Human Cytokine/Chemokine Discovery Assay Array (Eve Technologies, Calgary, AB) according to the manufacturer’s protocol.

### Fecal Short-Chain Fatty Acids (SCFAs)

SCFAs were extracted from 150 mg stool samples using acidified water and derivatization and were quantified using high-performance liquid chromatography with a diode array detector (HPLC-DAD), as previously described(73). Briefly, 100 uL of the extract was derivatized with 3-nitrophenylhydrazine hydrochloride in pyridine medium, resulting in the dissolution of SCFA-hydrazides in diethyl ether. Following evaporation, 20 uL of 50% methanol-dissolved SCFAs was injected into the HPLC-DAD. Analytes were separated on a C18 column at 40°C using a gradient of water and acetonitrile containing 0.05% trifluoroacetic acid (8% - 100%) at a flow rate of 0.8mL/min, with a 30-minute elution time. The absorbance of the SCFA-hydrazides was detected at a wavelength of 230nm.

### Fecal bacterial DNA extraction and sequencing

A total of 84 stool samples collected from 30 participants who completed the baseline assessment and at least one of the two follow-up physical function tests and sample collection at months 3 and 6 were selected for shotgun metagenomic sequencing. Genomic DNA of fecal bacteria was extracted from ∼500 mg stool samples using the FastDNA Spin Kit with bead beating (MP Biomedicals, Lachine, QC, Canada) for 40 s × 3 times. Purified DNA samples were quantified using Invitrogen Quant-iT™ PicoGreen™ dsDNA Assay Kit (Fisher Scientific, ON, Canada) and diluted to 50 ng/uL prior to sequencing. Shotgun metagenomic sequencing was conducted on a NovaSeq S2 300 cycle (2 × 150 bp, Centre for Health Genomics and Informatics, Calgary, AB, Canada) with NEBNext® Ultra™ II FS DNA Library Prep Kit (New England Biolabs, Ipswich, MA, USA) to generate 1250 GB of data (4100 M reads).

### Non-assembly based metagenomic composition and function profiling

Metagenomic shotgun sequencing reads were quality controlled using the KneadData pipeline (https://huttenhower.sph.harvard.edu/kneaddata). Briefly, adapter sequences and low-quality reads were trimmed and removed using Trimmomatic (v0.39)(75). Human genome contamination was aligned and discarded using Bowtie 2 (v2.5.4)(76). Species-level taxonomy of quality controlled reads was profiled using MetaPhlAn 4 (v4.1.1) by aligning shotgun reads with ∼ 5.1 M unique clade-specific maker genes, with the output presented as relative abundance (%)(23). Metagenomic gene families, metabolic pathways, and molecular functions were profiled using HUMAnN 3 (v3.9) with ChocoPhlAn 3 and UniRef90 databases(24). Output data was presented as Counts Per Million (CPM) by normalizing total counts of each category to the total reads per sample at both the community and species levels. Structural differences in the metagenomic compositional and functional profiles were assessed using the Shannon index for α-diversity and the Bray Curtis distance matrix for community dissimilarity (β-diversity). Non-metric multidimensional scaling (NMDS) was applied to visualize the dissimilarities in a two-dimensional space.

### Metagenomic assembly, binning, phylogenetic identification and abundance calculation

In addition to non-assembly-based compositional and functional profiling, quality-controlled reads were subjected to contig assembly with metaSPAdes (v3.15.5)(77). The assembled contigs were binned using MetaBAT2 (v2.17)(78) and MaxBin2 (v2.2.7)(79). The generated bins were quality controlled with CheckM2 (v1.1.0)(80) and dereplicated with dRep (v3.4.2)(81) to retain high-quality metagenomic assembled genomes (MAGs) with a completeness > 70% and contamination < 5%. Accurate taxonomic classification was assigned to MAGs by aligning against the Genome Taxonomy Database (GTDB) release 207 using GTDB-Tk (v2.2.6). The phylogenetic tree of MAGs was constructed using PhyloPhlAn 3.0 (v3.1.1) with multiple-sequence alignment algorithm MAFFT and tree reconstruction method RAxML (Randomized Axelerated Maximum Likelihood). The refined phylogenetic tree was visualized with iTOL (v7.1.1). Genome-wide abundance of MAGs was calculated by re-mapping the quality-controlled reads to each MAG and expressed as Transcript Per Million reads (TPM) using CoverM (v0.7.0)(82).

### Carbohydrate-active enzyme (CAZyme) annotation and substrate prediction

Sample-wide and MAG-wide microbial utilization of complex carbohydrate was characterized by annotating CAZymes and predicting their corresponding carbohydrate substrates at the CAZyme gene cluster (CGCs) level using dbCAN3 (25). First, gene models annotation was performed using Prokka (v1.14.5) (83) with assembled contigs (> 2 kbp) for sample-wide analysis and with MAGs for MAG-wide analysis. CAZymes were then annotated at the sub-family level, and corresponding CGCs were identified using run_dbcan (v4.1.4) (25, 84). Next, carbohydrate substrates were predicted at the CGC level using two approaches: 1) querying the dbCAN-PUL database (for both sample-wide and MAG-wide analyses), which contains experimentally characterized glycan substrates of Polysaccharides Utilization Loci (PULs); and 2) applying a majority voting approach across all CAZyme subfamilies with substrate prediction based on dbCAN-sub HMMdb annotations (for sample-wide analysis only)(25).

Following the CAZyme annotation and substrate prediction, for sample-wide analysis, reads were mapped back to the nucleotide coding sequences (CDS) of annotated contigs to calculate abundance of CAZymes, PULs, and CGCs, expressed as TPM. The abundance of PUL and CGC substrates per sample was determined by summing the abundance of PULs and CGCs with the same substrate predictions. For MAG-wide analysis, the metabolic potential of each MAG in utilizing specific substrates was determined by summing the number of PULs with the same substrate prediction. The potentials of MAGs in fructan and host glycan degradation were further ranked using the ‘*rank’* function in R, by excluding zero and missing values and assigning even ranks to even values, for visualization in iTOL(85).

### Determination of glycan-degrading enzyme activities in stool samples

The glycan-degrading enzyme activities of fecal bacteria toward host mucus- or dietary plant-derived glycans, namely sulfatase, β-N-acetylglucosaminidase, α-fucosidase, α-galactosidase, and β-glucosidase, in stool samples were determined as previously described (33, 86). Five 4-nitrophenyl-coupled substrates (potassium-4-nitrophenysulfate, 4-nitropheny N-acetyl-β-D-glucosamidine, 4-nitrophenyl-α-L-fucopyranoside, 4-nitrophenyl-α-D-galactopyranoside, and 4-nitrophenyl-β-D-glucopyranoside) and a serial dilution of 4-nitrophenol were used as chemical surrogates and standards for enzymatic activity detection, respectively. Following fecal protein extraction and concentration equalization, solubilized fecal protein was co-incubated with each of the five concentration-adjusted 4-nitrophenyl-coupled substrates for the appropriate amount of time (see details in Table 1). Upon incubation, enzyme catalyzes the hydrolysis of glycosidic/sulfate group bonds between the 4-nitrophenyl residue and releases free 4-nitrophenol, which can be detected by an increase in absorption at 405 nm.

**Table 1.**
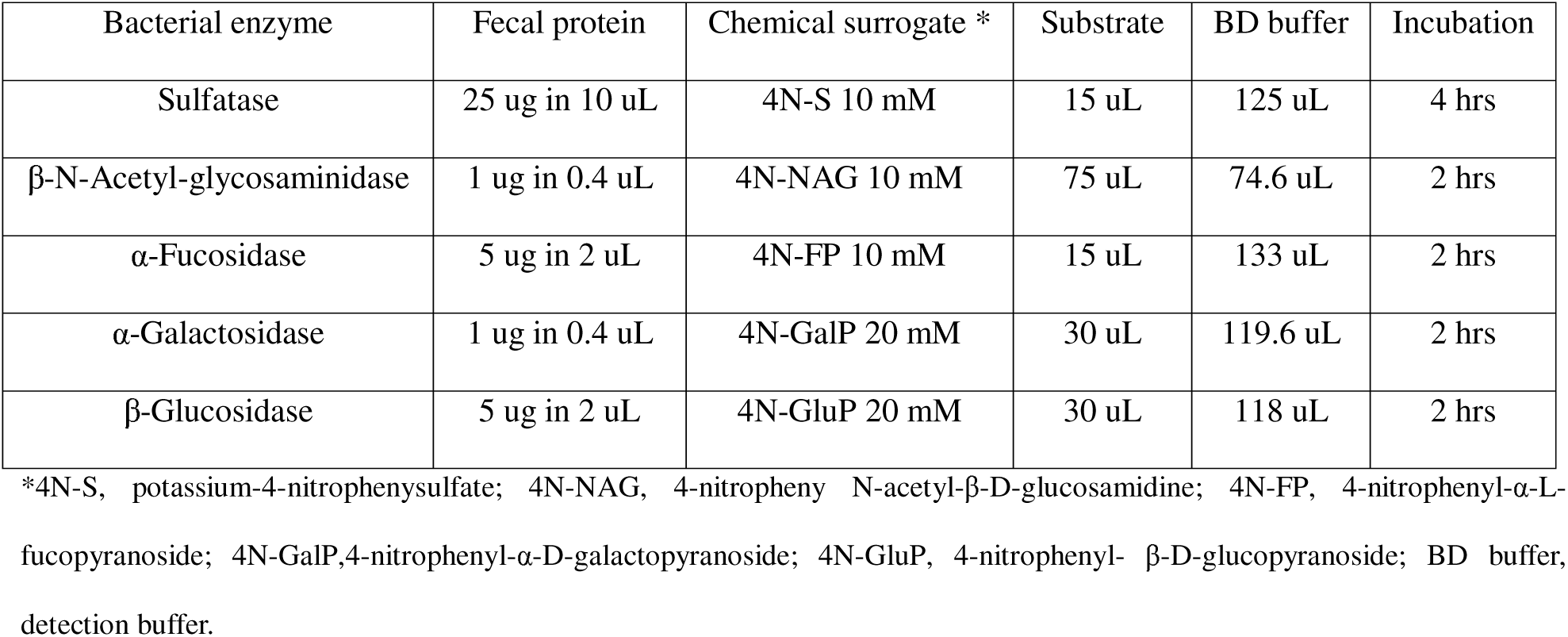
Reaction system (150 uL) and incubation time for enzymatic activity detection.

| Bacterial enzyme | Fecal protein | Chemical surrogate * | Substrate | BD buffer | Incubation |
| --- | --- | --- | --- | --- | --- |
| Sulfatase | 25 ug in 10 uL | 4N-S 10 mM | 15 uL | 125 uL | 4 hrs |
| β-N-Acetyl-glycosaminidase | 1 ug in 0.4 uL | 4N-NAG 10 mM | 75 uL | 74.6 uL | 2 hrs |
| α-Fucosidase | 5 ug in 2 uL | 4N-FP 10 mM | 15 uL | 133 uL | 2 hrs |
| α-Galactosidase | 1 ug in 0.4 uL | 4N-GalP 20 mM | 30 uL | 119.6 uL | 2 hrs |
| β-Glucosidase | 5 ug in 2 uL | 4N-GluP 20 mM | 30 uL | 118 uL | 2 hrs |

\*4N-S, potassium-4-nitrophenylsulfate; 4N-NAG, 4-nitrophenyl N-acetyl-β-D-glucosaminide; 4N-FP, 4-nitrophenyl-α-L-fucopyranoside; 4N-GalP, 4-nitrophenyl-α-D-galactopyranoside; 4N-GluP, 4-nitrophenyl-β-D-glucopyranoside; BD buffer, detection buffer.

Enzyme activities were presented in nkat/mg, which refers to the amount of cleaved substrates per second (mol/s, equal to kat) per milligram of solubilized fecal protein used in the assay. Briefly, five standard curves were created by replacing the same volume of fecal protein with serial diluted 4-nitrophenol in the same 150 uL reaction system for each of the five enzymes. The detected absorption of each sample or standard was plotted against time, from which the linear phase with the highest increase in absorption per unit time and containing 10-20 data points were selected for slope calculations (in Δ mAU/min). The log of the calculated slope values was then fitted with each liner standard curve to calculate the log of enzymatically released 4-nitrophenol mols. Finally, the log values were exponentiated, and the substrate turnover velocity was normalized by the amount of fecal protein used (in nmol/(min·µg)). The enzymatic activity was then expressed in nkat/mg by multiplying the velocity by 1000 (to convert ug to mg) and dividing by 60 (to convert minutes to seconds).

### Animal experiment

We re-analyzed a subset of data from a previous animal study examining the protective effect of prebiotics on knee joint health in a diet-induced obesity rat model(34). Briefly, 24 twelve-week-old male Sprague Dawley rats were randomly allocated to either a high-fat/high-sucrose diet (HFS; 20% fat, 50% sucrose, 20% protein, and 10% fiber/micronutrients (by weight); n=12) or HFS diet supplemented with 10% wt/wt oligofructose prebiotic (Orafti P95) (HFS+P) for 12 weeks. Serum samples were collected one week before the intervention (pre) and at endpoint (post), along with synovial fluid from the right knee joint, for cytokine profiling. Cytokines were quantified using a rat 27-Multiplex Discovery Assay (Eve Technologies, Calgary, AB) and we re-analyzed for those overlapping with human serum profiles, including IL-1β, IL-5, IL-4, IL-13, TNF, and IL-10. Absolute 16S rRNA gene copy numbers for total bacteria, *Akkermanisa muciniphila* and *Roseburia* spp. were quantified from 20 ng cecal bacterial genomic DNA using a specific primer set, as previously described(34).

The left knee was harvested, fixed in a 10% neutral buffered formalin, decalcified in Cal-Ex-II solution (Thermo-Fisher, Waltham, MA USA). The joint was then processed in a series of alcohols, infiltrated with paraffin, and embedded. Serial sagittal sections (10 µm thick) were cut, mounted on Fisher Superfrost Plus slides, and stained with hematoxylin, fast green, and safranin-O. Two independent, blinded observers scored the sections using the Modified Mankin score (range 0–24) for cartilage and OARSI for bone (range 0-12), where 0 indicates normal tissue and the highest score indicates the worst cartilage degeneration or bone changes(34, 87). Scores were recorded separately for the tibia and femur in both medial and lateral compartments (Medial Tibia, Medial Femur, Lateral Tibia, Lateral Femur). A total joint histologic score was calculated by summing the cartilage and bone scores. Synovium thickening and meniscus damage were scored on a 0–4 scale for each rat, as previously described (34, 87).

### Quantification And Statistical Analysis

#### Linear mixed model

For datasets following a normal distribution, including phenotypic data on body composition, physical function measures, α-diversity of metagenomic composition and function profiles, PUL profiles, serum cytokines (human and rat) and LPS, serum metabolome, fecal SCFAs, and CGC majority-voted substrate predictions, statistical differences were assessed using a linear mixed model. The model was fitted using the ‘*lme4’* package in R (v4.4.1), with group (prebiotic vs. placebo) and time (months 0, 3, and 6 or pre vs post) as fixed effects and participants or rats as a random effect. Post-hoc comparisons were performed using the ‘*emmeans’* package. A significant difference was defined as an adjusted *p* value < 0.05, with p-value adjustment using the Bonferroni, Benjamini-Hochberg, or Tukey correction procedure.

#### Non-parametric Statistical Analysis

For non-normally distributed datasets, including cecal bacterial abundance and the *A. muciniphila* /*Roseburia* spp. ratio in rat, statistical differences were compared using Kruskal-Wallis rank-sum test in using the ‘kruskal.test’ function R (v4.4.1).

#### Permutational Multivariate Analysis of Variance (PERMANOVA)

PERMANOVA was performed to assess the statistical significance of group, time, group × time, and individuality in explaining the Bray-Curtis distance matrix based dissimilarity of MetaPhlAn species, HUMAnN3 unstratified pathways, and dbCAN3 PULs. The ‘*adonis2*’function from the ‘*vegan’* package in R was used for the PERMANOVA analysis, with 999 permutations.

#### Random Forest Regression model

Phenotype-predictive random forest regression modes were trained using z-score transformed numeric variables from MetaPhlAn4 species, HUMAnN3 unstratified pathway, dbCAN3 PUL prediction, either individually or in combination with serum cytokine and serum metabolome as predictors. The models were trained using the ‘*train’* function from the ‘*caret’* package (88) in R, with bootstrapping set to 100 for model validation via the ‘*trainControl’* function. Model tuning was conducted by adjusting the ‘*mtry’* parameter, which controls the number of variables considered at each split. The tuning grid explored ‘*mtry’* values ranging from 12 to 48, with a step size of 2. Model performance was assessed using the coefficient of determination (R²) and Mean Absolute Error (MAE). R² measures the proportion of dependent variables explained by the model, while MAE calculates the average absolute difference between predicted and actual values.

#### Bacteria-Cytokine Co-occurrence Network Analysis

Bacteria-cytokine co-occurrence networks were constructed for prebiotic and placebo participants, using nine serum cytokines and MetaPhlAn4 species that showed significant correlations (FDR < 0.05, R^2^ > 0.45) with at least one cytokine as nodes. The network file was generated using the ‘*graph_from_data_frame’* function from the ‘*igraph’* package in R and visualized with Gephi (89) using the ‘*Fruchterman Reingold’* layout. Betweenness centrality of bacterial and cytokine nodes was calculated using the *betweenness* function from the ‘*igraph’* package (https://igraph.org/) to assess their role as bridges within the network.

#### Multi-Omics Factorial Analysis 2 (MOFA2) model

Prior to model training, datasets including MetaPhlAn4 species, HUMAnN3 unstratified pathway, dbCAN3 PUL predictions, serum cytokines, serum metabolome, and CGC majority-voted substrates were preprocessed by centering and scaling using the ‘*caret::preProcess’* function in R. The MOFA2 model was then constructed and trained using the ‘*MOFA2’* (90) package in R. The group- and time-aware settings were enabled through the ‘*MEFSTO’* (91) function to account for the experimental design. The model was trained with 5,000 iterations to ensure convergence, facilitating optimal factor extraction and robust data integration across multiple omics views. The ‘*get_variance_explained’* function from the ‘*MOFA2’* package was employed for variance decomposition. The ‘*get_weights’* function was used for factor weights extraction for each feature across views. The ‘*correlate_factors_with_coavriates’* function was utilized to generate correlation matrixes between the extracted latent factors and the outcomes of interest.

## DECLARATIONS

## Ethics approval and consent to participate

Ethical approval for the human study was provided by the Conjoint Health Research Ethics Board of the University of Calgary (REB17-2363) and was prospectively registered at ClinicalTrial.gov (NCT04172688). Ethical approval for the animal study was provided by the University of Calgary Life and Environmental Sciences Animal Care Committee.

## Consent for Publication

Not applicable.

## Availability of data and materials

Supplemental Tables S1–S16 provide details on post-intervention outcomes. Tables S1–S14 present results from human participants, covering physical function and body composition (Table S1), fecal microbiome composition and function (Tables S2–S3, S6–S9, S11–S14), as well as serum cytokines, LPS, and metabolome data (Tables S5 and S10). Tables S15–S16 present findings from the diet-induced obesity rat model, including serum/synovial cytokines (Table S15) and joint integrity scores (Table S16). The fecal shotgun metagenomic sequence data have been deposited in the NCBI Short Read Archive under Bioproject accession PRJNA1272060. The R scripts and datasets used for data analysis and visualization in both human and rat model are publicly available on the GitHub repository: https://github.com/Weilan2011/Osteoarthritis-Prebiotic-Multiomics. Upon request, the lead contact Dr. Raylene Reimer and technical contact Dr. Weilan Wang can provide any additional information required to reanalyze the reported data in this paper.

## Acknowledgements

The authors would like to acknowledge Dr. Rachel Schachar and Dr. Kelly Johnston for assistance with participant recruitment. Metabolomics data were acquired by Marija Drikic at the Calgary Metabolomics Research Facility (CMRF), which is supported by the International Microbiome Centre and the Canada Foundation for Innovation. This work was supported by a Weston Foundation Microbiome Initiative Grant and a McCaig Encore Catalyst Grant. WW was supported by an Eyes High Postdoctoral Fellowship. RF was supported by an Alberta Innovates Postdoctoral Fellowship and a Dr. Cy Frank Trainee Award in Nutrition, Bone and Joint Health. EWNT was supported by an Alberta Innovates Postdoctoral Fellowship.

## Authors’ Contributions

WW, RF, RAR, DAH, and KAS conceived the study; WW conducted the enzymatic analysis, gut microbiome analysis in humans and rats, and all other bioinformatics analysis; RF conducted the randomized controlled trial; SM conducted the metabolomics analysis; CM assisted with bioinformatics; EWNT conducted the SCFA analysis; RAS, JLR, NA, and WH conducted the rat joint histology study and analysis; EVM conducted the self-reported data analysis; WW, RAR and KAS drafted the manuscript; RAR had primary responsibility for the study; all authors reviewed and approved the final submitted manuscript.

