## Supplemental Figures for "Microbial mechanisms underlying prebiotic-linked improvements in physical function and metabolism in knee osteoarthritis and obesity"

**Competing Interests**

WW, RF, SM, RAS, CM, JLR, NA, EVM, EWNT, DAH, KAS, and WH declare no competing interests. RAR has received speaker honoraria from Beneo for work distinct from the current study.

**Supplemental Figures**

**Figure S1. Integration of prebiotic-responsive MetaPhlAn4 species with phenotypic outcomes**

**(A)** MetaPhlAn4 species significantly responsive to prebiotic intervention (FDR corrected q < 0.05), identified by using MaAsLin2. **(B)** Performance of random forest model in predicting phenotypic outcomes using MetaPhlAn4 species-level taxonomy, HUMAnN3 metabolic pathway, dbCAN3 PUL, individually or in combination with serum cytokines and metabolites as predictors. R² (coefficient of determination) represents the proportion of variance explained by the predictors. **(C)** Representative MetaPhlAn4 species associated with trunk fat (%), total lean, and hand grip strength. Only the 15 species with significant Spearman’s correlations (*p* < 0.05) and the greatest coefficients are shown. Positive (+) and negative (-) signs within the panels indicate species significantly increased or decreased (FDR < 0.1) in prebiotic participants, as determined by MaAsLin2.

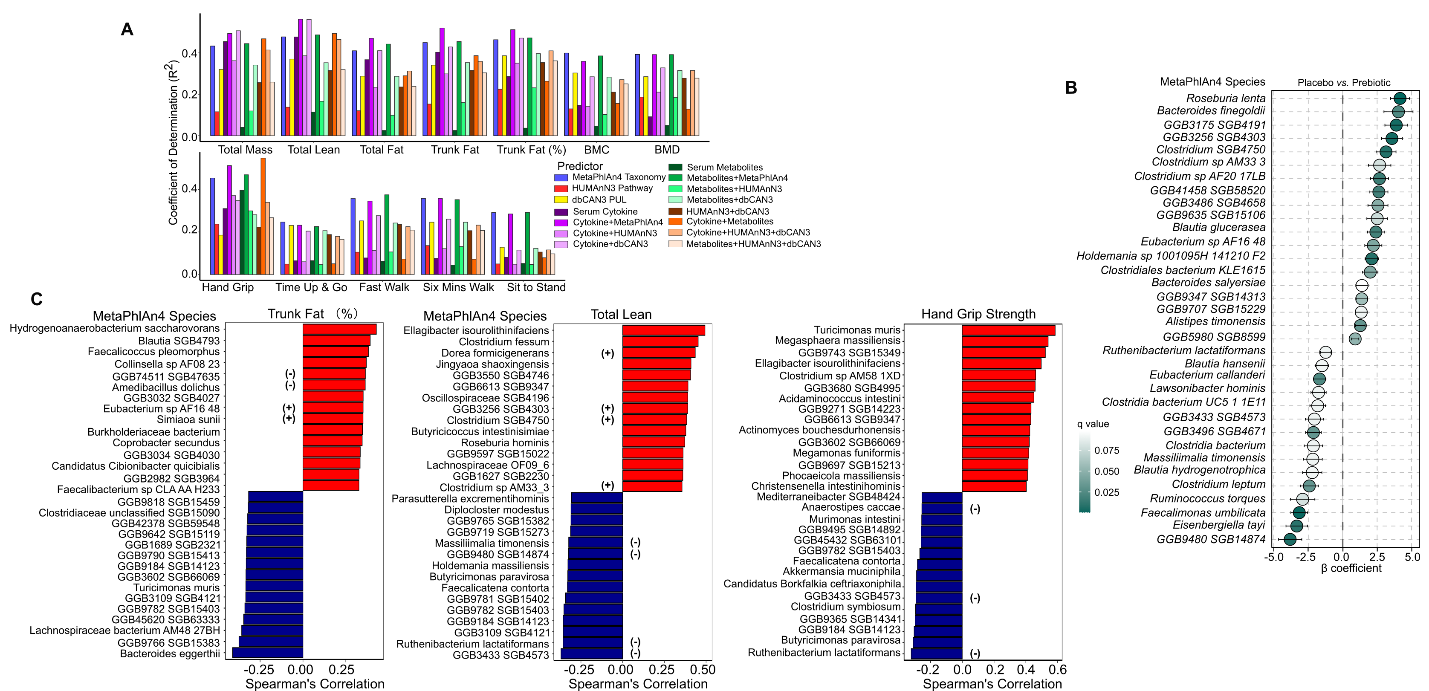

**Figure S2. Longitudinal profiles of sample-wide carbohydrate feedstock utilization**

Sample-wide carbohydrate feedstock utilization was quantified by summing the abundance of CGCs (CAZyme Gene Cluster) with the same predicted substrate, applying a majority voting approach across all CAZyme subfamilies per CGC. Data was represented as TPM.

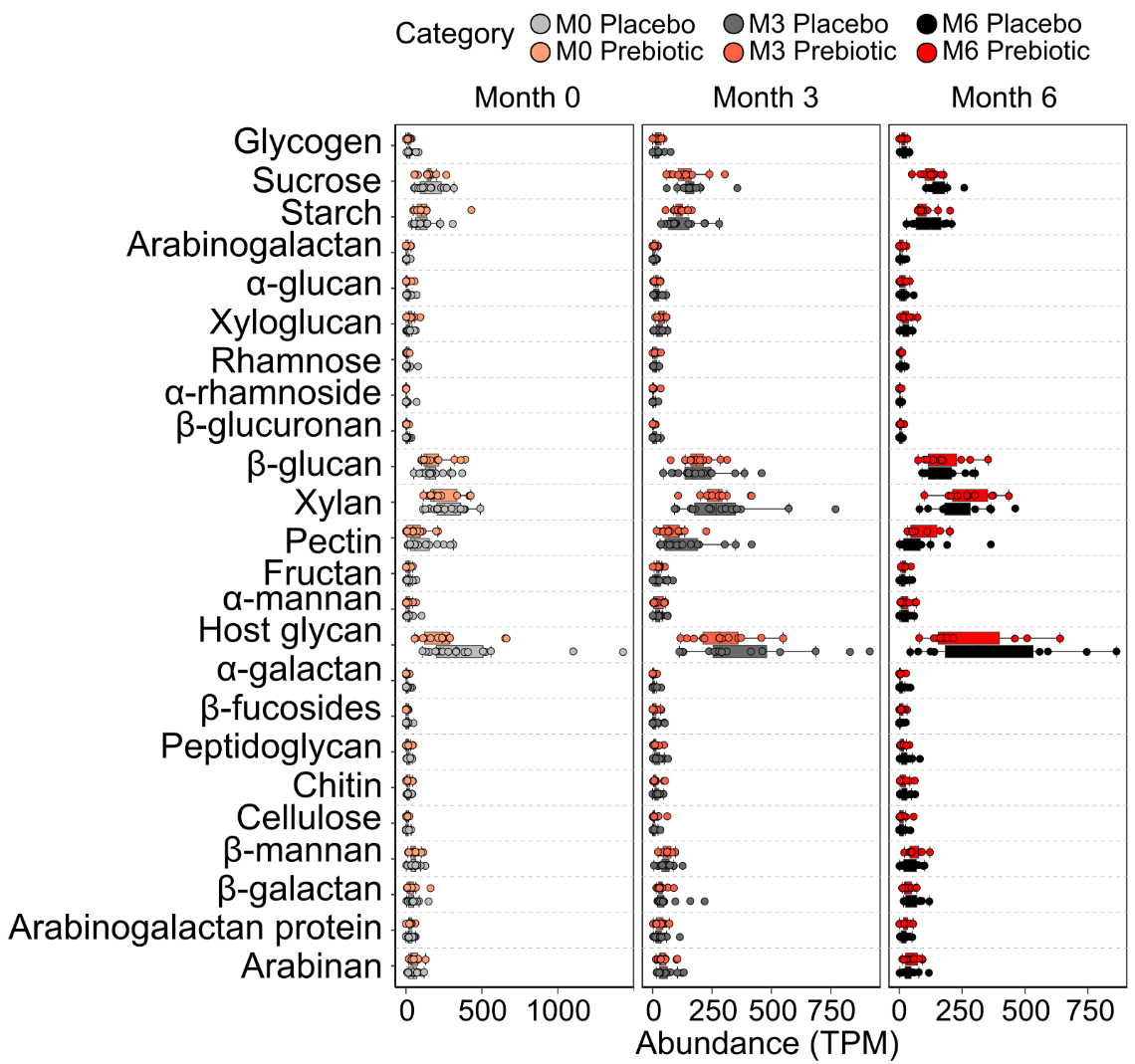
